# Victimisation, depression and suicidal ideation: understanding interlinkages among youth in North India through structural equation modelling

**DOI:** 10.64898/2026.09.14.26362969

**Authors:** Adhish Kumar Sethi, PVM Lakshmi, Vikas Kumar Bhatia, Shubh Mohan Singh

**Affiliations:** Department of Community Medicine and School of Public Health Postgraduate Institute of Medical Education and Research, Chandigarh, India; Department of Psychiatry Postgraduate Institute of Medical Education and Research, Chandigarh, India

**Keywords:** depression, suicide, violence, youth, victimisation

## Abstract

**Background:** Suicide and violence are major contributors to youth mortality. Effective prevention strategies require understanding of their psychological underpinnings. We therefore aimed to determine the interconnections among victimisation, physical violence, depression and suicidal ideation and attempts, among young adults in a North India city.

**Methods:** We used data from a survey of young adults aged 18–22 years, recruited across six colleges by random sampling. Our theory-based structural equation model specified suicidal ideation and attempts as an outcome of victimisation, indicated by dating violence, campus bullying and cyberbullying. We coded ‘suicidal ideation and attempts’ as a single ordinal variable, with five levels from ‘did not consider suicide’ to ‘had a suicide attempt requiring medical treatment’. We used the probit link function and theta parameterisation for categorical endogenous variables. We computed standardised path coefficients, p-values and global fit measures.

**Results:** Among 752 survey respondents (median age 19 years, 64.4% women), 16.6% had been involved in a physical fight, 24.2% had had depressive symptoms and 4.5% had seriously considered suicide in the past 12 months. We found indirect associations between victimisation and suicidal ideation and attempts, mediated by physical violence and depressive symptoms. Direct effects of victimisation on suicidal ideation and attempts were negligible. Global model fit was acceptable.

**Discussion:** Our findings suggest that youth victimisation predisposes to suicidal behaviour, by increasing physical aggression and depression. Prevention of suicide among youth requires multi-level action, including measures to stop bullying and violence, life skills education, and treatment of underlying depression.

## Introduction

### Mental health in youth

The period from 15–24 years of age is an important period in an individual’s life. It is a period of transition from childhood to adulthood, whereby one becomes a fully functioning member of society. Though terminology varies, most authors label this as the period of “youth” (Patton et al., 2016). The profound changes in body structure, emotional functioning and societal roles that characterise youth are also accompanied by increased vulnerability to mental health issues. Adolescence and young adulthood often mark the onset of mental disorders such as mood disorders. Global literature suggests that the incidence of mental illness peaks around 15 years of age, and the median age of onset is 19–20 years (McGrath et al., 2023). Aggression and suicide are also important behavioural concerns among youth (Lee et al., 2022). Within the 15–19 years age group, older adolescents are more vulnerable to suicide (Hughes et al., 2023).

Together, these mental disorders create an enormous health impact worldwide. It is estimated that 4% of the disability-adjusted life years (DALYs) among persons aged 15–29 years globally are due to mood disorders (World Health Organization, 2020a). Most of this burden is due to depression. Further, aggressive behaviours and violence are responsible for about 9% of deaths among persons aged 15–29 years globally. Suicide is responsible for 8% of the deaths in this age group (World Health Organization, 2020b).

### Youth mental health: the Indian scenario

A fifth of the world’s young people live in India (McFarlane, 2023). The specific mental health needs of youth are increasingly being recognised in India (Ministry of Health and Family Welfare, Government of India, 2014; Sahadevan et al., 2023). The National Mental Health Survey conducted in 12 states of India from 2015 to 2016 found that 3% of persons aged 18–29 years had depressive disorders, and 0.9% were at high risk of suicide (Gururaj et al., 2016). Interpersonal violence contributes to 3% of deaths in the 15–29 years age group in India (World Health Organization, 2020b). Data on reported crime, from the National Crime Records Bureau in India, show that from 2017 to 2022, the annual number of murder victims in the 18–30 years age group declined by about 2%. In contrast, the annual number of rapes reported by women in this age group rose by nearly 25% during this period (National Crime Records Bureau, 2019, 2023).

The changing socio-economic and cultural milieu of India means that more youth are now availing higher education (International Institute for Population Sciences (IIPS) & ICF, 2022). Youth in higher educational institutions in India are exposed to various risk factors for emotional and behavioural problems, including academic pressures, financial difficulties, and bullying (Bhola et al., 2016; Chaudhary et al., 2023). At the same time, experience from other countries has shown that universities can be a medium for imparting life skills education and reducing suicide risk among youth (Harrod et al., 2014). Thus, college-going youth are an important group for mental health initiatives in India.

### The role of victimisation

The consensus view in the literature is that youth aggressive behaviour, depression and suicide are complex phenomena. Their causation is multifactorial, covering biological, psychological and socio-environmental domains. Within the psychosocial domain, previous literature has identified victimisation as a predisposing factor for aggression, depressive symptoms and suicidal ideation among youth (Cruz-Manrique et al., 2021; Ortiz-Marcos et al., 2022). Victimisation among youth is a broad construct, and takes different forms in different cultural contexts. These include bullying, physical violence, and sexual abuse.

Bullying in adolescence has shown positive associations with mental disorders (R. Bhatia, 2023) and suicide (Hasan et al., 2021), while the association between sexual abuse and mental disorders is less consistent (McKay et al., 2021). Violence during dating is a known risk factor for suicide attempts among young women (Miranda-Mendizabal et al., 2019). One early psychological model that explains the link between adverse life experiences, such as victimisation, and adverse mental health outcomes, such as suicide, is that of internalisation. In this model, the adverse experience generates hostile feelings in the individual. In a maladaptive process, these hostile feelings are turned inwards, which results in hopelessness and suicidal ideation (Lester et al., 1979; Shneidman, 1967). Victimisation among youth may also lead to increased aggression (Camacho et al., 2021). In this context, aggression has been seen as a failure of normal coping mechanisms, or as a defensive measure to display and maintain social status (Camacho et al., 2021; Liu et al., 2013).

### Methodological approaches

Understanding these interrelationships can help develop effective interventions for prevention of violence and suicide in schools and colleges (Mytton et al., 2006; Sutter et al., 2023). In this regard, many investigators have attempted to quantify the associations of victimisation, aggression and suicide in youth. Some have used cross-sectional data to derive simple measures of association between victimisation and violence perpetration (Herrenkohl & Jung, 2016; Jeong et al., 2015), or victimisation and suicide (Biswas et al., 2020). While these studies provide useful insights, they do not elucidate intermediates in the causal path from victimisation to the outcomes of interest, and do not show temporal relationships. With availability of powerful computers and the growth of advanced statistical techniques, behavioural scientists have increasingly turned to structural equation modelling for understanding complex relationships between constructs. The advantage of this technique is that it simultaneously considers the variances and covariances of multiple variables of interest, allowing quantification of direct and indirect effects (Kline, 2023). As an example, a structural equation modelling study among high-school students in the Netherlands found that victimisation was positively associated with depression not just directly, but also by its negative effects on personal identity, which protects against depression (Van Hoof et al., 2008). A further improvement comes from cross-lagged models, which extend structural equation modelling to longitudinal studies. Use of such models has revealed reciprocal relationships between victimisation and mental disorders among youth, each leading to the other at subsequent time points (Le et al., 2019; Zhu et al., 2022).

### Literature from India

While these issues have been discussed in the global literature, few studies have looked at interrelationships between victimisation and adverse mental health outcomes among youth specifically in the cultural context of India. A cross-sectional study conducted among adolescents in seven countries, including India, during the COVID–19 pandemic found that victimisation was positively associated with stress and depression, with resilience mediating its effect (Bravo-Sanzana et al., 2023). Results from the Understanding the Lives of Adolescents and Young Adults (UDAYA) study indicated that victims of cyber-bullying were more likely to suffer from depressive symptoms and suicidal ideation, while self-efficacy and parental communication played a protective role (Maurya et al., 2022, 2023). Considering that the sampling in these studies focused on specific age groups in selected regions of India, the extent to which their results can be generalised to youth across India is unclear. Understanding is particularly lacking for risk factors of violent and aggressive behaviours among youth in India. A study conducted among adolescents aged 15–19 years in Karnataka, India, found that persons victimised by violent activities were more likely to engage in violent behaviour (Swain et al., 2014). Again, given the diversity of cultures within India, the generalisability of this result to the broader Indian population is unclear.

Overall, the available studies provide limited understanding of the pathways linking victimisation to aggression and suicidal behaviour among Indian youth. Context-specific knowledge of such linkages will help design locally relevant prevention programmes for depression, suicide and violence, so that the burden of death and disability among youth may be reduced. Such knowledge will be especially useful for government programmes and non-governmental organisations which are working for youth mental health in India (LonePack, c2024; Ministry of Health and Family Welfare, Government of India, 2015).

### Current study

Therefore, we aimed to quantify the interlinkages among victimisation, aggression, depression and suicidal ideation and attempts, among college-attending youth in Chandigarh, India. We used data from a representative survey conducted in 2018. Chandigarh, a prominent site for higher education in northern India, has nearly 30 government and private colleges enrolling students from multiple regions of India (Department of Education, Chandigarh Administration, c2017; Panjab University, c2024). We could not find any previous studies on this topic from this setting.

## Methods

### Study setting, design and population

This was a secondary analysis of cross-sectional data collected in 2018 (V. K. Bhatia et al., 2026). Participants were students aged 18–22 years, enrolled in colleges in Chandigarh.

### Sampling and study size

Sampling and study size calculation are described in detail elsewhere (V. K. Bhatia et al., 2026). Briefly, we selected participants by multistage stratified random sampling across six colleges (about 150 students approached per college). The primary objective of the study was to assess the prevalence and clustering of behavioural risks among youth, including injury risks, suicidal ideation, dietary risks, sexual risks and substance use. For that objective, a study size of 793 individuals was sufficient. We were able to recruit 752 participants for the study.

To judge the adequacy of this study size for the present analysis, involving structural equation modelling, we used a rule of thumb of at least 200 individuals, and at least 20 per free parameter to be estimated (Jackson, 2003). Our initially hypothesised model (Supplementary Figure 1) had 31 free parameters, which meant data from 620 individuals were required. Therefore, we considered our study size adequate.

### Study tool and data collection

During the survey, we used the Centers for Disease Control and Prevention (CDC) Youth Risk Behaviour Surveillance System (YRBSS) 2017 Standard High School questionnaire to collect data (Centers for Disease Control and Prevention, 2023). It was an English, self-administered, paper-based, structured questionnaire. To reduce questionnaire length, we removed the items about carrying or using weapons in college, as we expected these behaviours to be extremely rare in the Indian context. We did not make any changes in the other questions on victimisation, physical fights, suicidal ideation or depressive symptoms. The questionnaire was pre-tested among 10 students before use. It included 74 items covering various behavioural risks ((V. K. Bhatia et al., 2026)), and required 30–45 minutes to complete. To maintain privacy, the questionnaire did not ask students for their names. Students completed the questionnaires in classrooms, under the supervision of one of the investigators.

### Data processing and preparation

We performed data processing and analysis with R version 4.3.0. The items considered in the present analysis covered experiences over the 12 months preceding the survey, regarding sexual abuse, physical violence while dating, offline and cyber-bullying, depressive symptoms lasting at least two weeks, suicidal ideation and attempts, and involvement in physical fights. We also included age and gender as model covariates.

We initially cleaned data to remove logical inconsistencies in responses (e.g., a participant reporting no suicide attempt, but reporting a suicide attempt requiring medical treatment). In such cases, we set both responses to missing. Then, we excluded individuals with valid data for <20 of the 74 items. We recoded all variables as binary (yes/no), except for age, gender and suicide. We combined data from the four items on suicidal ideation and attempts in the last 12 months, into a single, five-level ordinal variable: ‘did not consider suicide’, ‘considered but did not plan suicide’, ‘considered and planned but did not attempt suicide’, ‘attempted suicide but did not require medical treatment’, and ‘had a suicide attempt requiring medical treatment’.

### Examining distributions

We examined the distributions of the responses, in terms of counts and proportions for categorical variables, and median and interquartile range (IQR) for age. We also examined bivariate distributions by cross-tabulation. We calculated polychoric correlation for pairs of ordered categorical variables, and polyserial correlations for age and categorical variables. These correlations accounted for the categorical nature of the variables, by assuming underlying normally distributed variables which generate the observed categories (Holgado– Tello et al., 2010).

### Examining factor structure of the ‘victimisation’ construct

We performed exploratory factor analysis for the five binary variables pertaining to victimisation, using geomin rotation. We examined the factor loadings, unique and common variances, eigenvalues and global fit indices for one-factor and two-factor model. However, with these variables, we observed Heywood cases with negative unique variance for the sexual abuse variables. Considering the large correlation between ‘sexual abuse’ and ‘sexual abuse during dating’, we decided to drop ‘sexual abuse during dating’ and tested a one-factor model for the remaining four variables.

### Model specification

We developed a preliminary model based on a review of the literature (Supplementary Figure 1) (Blain-Arcaro & Vaillancourt, 2017; Jun et al., 2015; Maurya et al., 2022). Considering the large correlation between ‘physical fight’ and ‘physical fight on campus’, we dropped ‘aggression’ as a latent variable, and kept only ‘physical fight’ in the model. Since ‘sexual abuse’ had a small loading on the victimisation factor, we included it as a separate variable uncorrelated with victimisation, rather than an indicator of victimisation. Thus, our specified model had nine observed variables, one latent variable, and 30 free parameters (Figure 1a).

**Figure 1:**
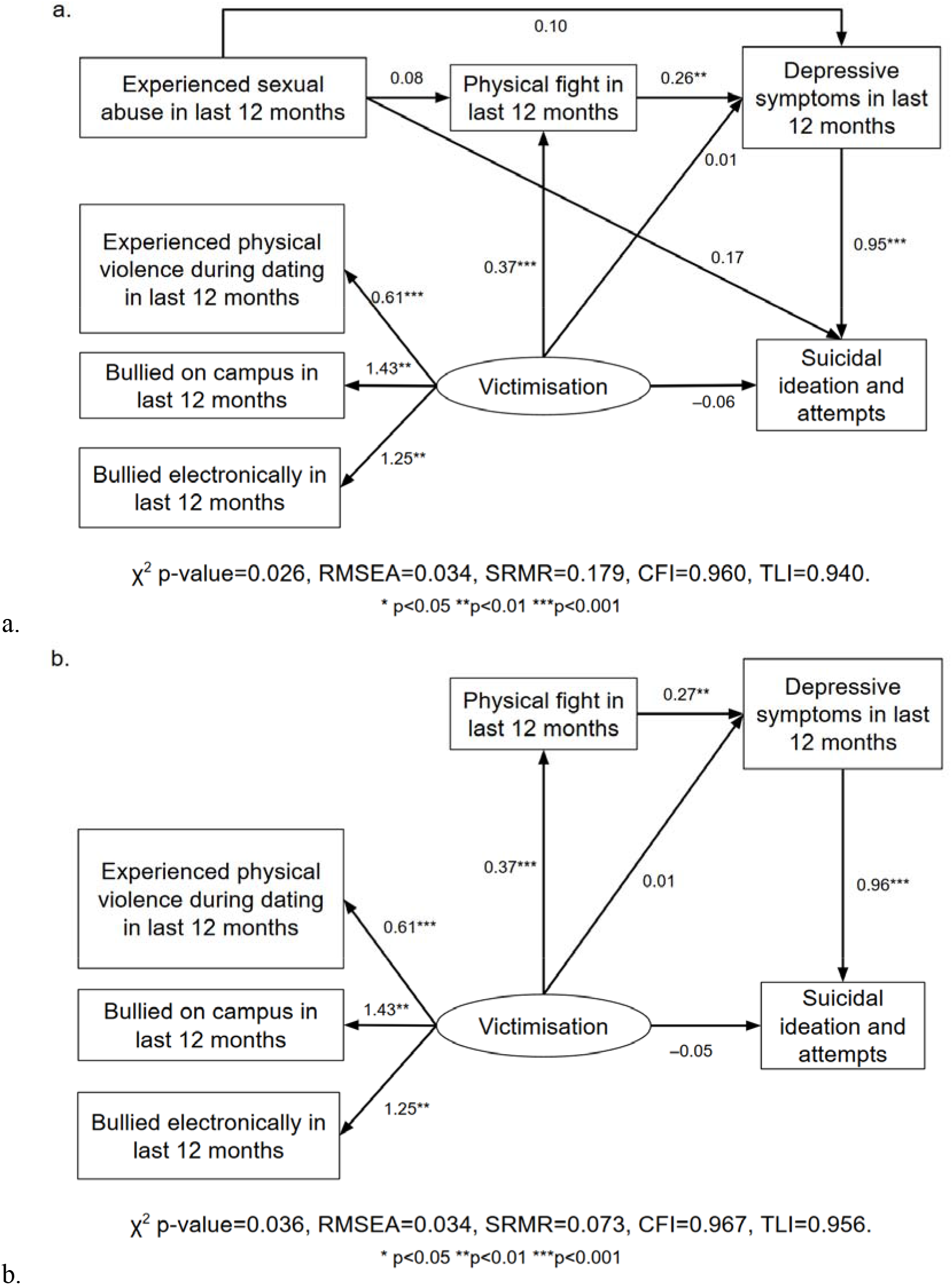
Structural equation models for victimisation, aggression, depression and suicidal ideation and attempts among young adults attending colleges in Chandigarh. a. With sexual abuse included in the model, b. With sexual abuse dropped from the model. Notes: Path coefficients were adjusted for age and gender (adjustment paths not shown in the diagrams for clarity), and standardised for the latent variable ‘victimisation’. Categorical endogenous variables had unit residual variance, under the theta parameterisation. Both models used data from 752 individuals.

### Model fitting and assessment

We fitted the model using the diagonally weighted least squares estimator, and the probit link for ordered categorical variables (e.g., cyber-bullying, suicide). The probit link meant that each categorical variable was described by an underlying normally distributed variable, and a change of category corresponded to the normally distributed variable crossing a particular threshold (Rosseel et al., 2024). We used theta parameterisation, which scaled categorical variables to unit variance. The fitting algorithm accounted for missing data, considering all individuals who provided data for a particular variable for mean, variance and covariance calculation. Thus, the only excluded individuals were those who had missing data for an exogenous variable, and those who had missing data for all endogenous variables. For the fitted model, we reported path coefficients, confidence intervals (CIs) for the path coefficients, and p-values, standardised for unit variance of the ‘victimisation’ latent variable. We assessed global model fit using the chi-squared test for goodness of fit, root mean squared error of approximation (RMSEA), standardised root mean squared residual (SRMR), comparative fit index (CFI) and Tucker–Lewis index (TLI).

Considering that sexual abuse was quite rare in our sample (<5%), and had weak and uncertain associations with the endogenous variables, we also examined the effect of dropping this variable on path coefficients and model fit (Figure 1b).

### Ethical considerations

The Institute Ethics Committee of the Postgraduate Institute of Medical Education and Research, Chandigarh approved the study protocol (letter number INT/IEC/2018/000791, dated May 24, 2018). Students provided informed, written consent for participation. Investigators ensured that data were kept private and confidential.

## Results

### Participant characteristics

Of 995 eligible students invited, 752 (75.6%) returned completed questionnaires. Table 1 summarises the characteristics of study participants. The majority (64.4%) identified as women. The median age of participants was 19 years. Whereas 2% had faced sexual abuse in the past 12 months, 4% had been physically hurt by a person they were dating, 10.2% had been bullied, and 10.8% had been cyber-bullied. Nearly 17% had been involved in a physical fight in the preceding 12 months. Almost a fourth of participants reported depressive symptoms for two weeks or more in the past 12 months, and 4.5% had seriously considered suicide in the past 12 months. Three individuals (0.4%) had attempted suicide in the past 12 months. Missing data were <5% for all variables. None of the participants had missing data for age or gender, and none had missing data for all endogenous variables in the model.

**Table 1:** Demographic characteristics, victimisation, physical fights and mental health among study participants.

| Characteristic | Men (n=268) | Women (n=484) | Total (n=752) |
| --- | --- | --- | --- |
| **Age in completed years: median (IQR)** | 19 (19, 20) | 20 (19, 21) | 19 (19, 21) |
| **Experienced sexual abuse in last 12 months** |  |  |  |
| Yes | 6 (2.2) | 9 (1.9) | 15 (2.0) |
| *Once* | 4 | 6 | 10 |
| *2 or 3 times* | 2 | 2 | 4 |
| *4 or 5 times* | 0 | 1 | 1 |
| *6 or more times* | 0 | 0 | 0 |
| No | 252 (94.0) | 462 (95.5) | 714 (94.9) |
| Missing data | 10 (3.7) | 13 (2.7) | 23 (3.1) |
| **Experienced sexual abuse during dating in last 12 months** |  |  |  |
| Yes | 4 (1.5) | 8 (1.7) | 12 (1.6) |
| *Once* | 3 | 6 | 9 |
| *2 or 3 times* | 1 | 2 | 3 |
| *4 or 5 times* | 0 | 0 | 0 |
| *6 or more times* | 0 | 0 | 0 |
| No, or did not date in last 12 months | 254 (94.8) | 463 (95.7) | 717 (95.3) |
| Missing data | 10 (3.7) | 13 (2.7) | 23 (3.1) |
| **Experienced physical violence during dating in last 12 months** |  |  |  |
| Yes | 18 (6.7) | 11 (2.3) | 29 (3.9) |
| *Once* | 13 | 7 | 20 |
| *2 or 3 times* | 3 | 1 | 4 |
| *4 or 5 times* | 1 | 2 | 3 |
| *6 or more times* | 1 | 1 | 2 |
| No, or did not date in last 12 months | 240 (89.6) | 460 (95.0) | 700 (93.1) |
| Missing data | 10 (3.7) | 13 (2.7) | 23 (3.1) |
| **Bullied on campus in last 12 months** |  |  |  |
| Yes | 37 (13.8) | 40 (8.3) | 77 (10.2) |
| No | 231 (86.2) | 444 (91.7) | 675 (89.8) |
| **Bullied electronically in last 12 months** |  |  |  |
| Yes | 35 (13.1) | 46 (9.5) | 81 (10.8) |
| No | 233 (86.9) | 438 (90.5) | 671 (89.2) |
| **Involved in physical fight in last 12 months** |  |  |  |
| Yes | 54 (20.1) | 71 (14.7) | 125 (16.6) |
| *Once* | 24 | 31 | 55 |
| *2 or 3 times* | 13 | 17 | 30 |
| *4 or 5 times* | 10 | 10 | 20 |
| *6 or 7 times* | 2 | 3 | 5 |
| *8 or 9 times* | 0 | 0 | 0 |
| *10 or 11 times* | 0 | 0 | 0 |
| *12 or more times* | 5 | 10 | 15 |
| No | 208 (77.6) | 412 (85.1) | 620 (82.4) |
| Missing data | 6 (2.2) | 1 (0.2) | 7 (0.9) |
| **Involved in physical fight on campus in last 12 months** |  |  |  |
| Yes | 11 (4.1) | 8 (1.7) | 19 (2.5) |
| *Once* | 6 | 6 | 12 |
| *2 or 3 times* | 2 | 1 | 3 |
| *4 or 5 times* | 0 | 0 | 0 |
| *6 or 7 times* | *0* | *1* | *1* |
| *8 or 9 times* | *0* | *0* | *0* |
| *10 or 11 times* | *0* | *0* | *0* |
| *12 or more times* | *3* | *0* | *3* |
| No | 251 (93.7) | 475 (98.1) | 726 (96.5) |
| Missing data | 6 (2.2) | 1 (0.2) | 7 (0.9) |
| **Depressive symptoms in last 12 months** |  |  |  |
| Yes | 75 (28.0) | 107 (22.1) | 182 (24.2) |
| No | 193 (72.0) | 377 (77.9) | 570 (75.8) |
| **Suicidal ideation and attempts** |  |  |  |
| Did not consider suicide | 251 (93.7) | 445 (91.9) | 696 (92.6) |
| Considered but did not plan suicide | 5 (1.9) | 17 (3.5) | 22 (2.9) |
| Considered and planned but did not attempt suicide | 3 (1.1) | 5 (1.0) | 8 (1.1) |
| Attempted suicide but did not require medical treatment | 0 (0.0) | 0 (0.0) | 0 (0.0) |
| Had a suicide attempt requiring medical treatment | 1 (0.4) | 2 (0.4) | 3 (0.4) |
| Missing data | 8 (3.0) | 15 (3.1) | 23 (3.1) |

Data are presented as count (percentage), unless specified otherwise. Percentages within columns add to 100%.

The correlation matrix of the study variables is shown in Supplementary Appendix 1. The polychoric correlation for ‘sexual abuse’ and ‘sexual abuse during dating’ was 0.993, and that for ‘physical fight’ and ‘physical fight on campus’ was 0.871. All other correlations were <0.8 in absolute value.

### Factor analysis

The results of factor analysis for the four-indicator, one-factor model for victimisation are shown in Table 2. The factor accounted for 36.6% of the total variance. The global fit measures were: chi-squared p-value 0.113, RMSEA=0.040, SRMR=0.113, CFI=0.975 and TLI=0.925.

**Table 2:** Factor analysis of the hypothesised victimisation indicators, for individuals who had complete data for all four variables (n=729)

| **Variable** | **Standardised factor loading** | **p-value for standardised factor loading** | **Unique variance** | **Communality** |
| --- | --- | --- | --- | --- |
| Experienced physical violence during dating in last 12 months | 0.426 | 0.001 | 0.818 | 0.182 |
| Experienced sexual abuse in last 12 months | -0.099 | 0.480 | 0.990 | 0.010 |
| Bullied on campus in last 12 months | 0.914 | <0.001 | 0.164 | 0.836 |
| Bullied electronically in last 12 months | 0.662 | <0.001 | 0.561 | 0.439 |

### Structural equation modelling

The results of structural equation modelling are shown in Figure 1 and Table 3. The model with ‘sexual abuse’ showed that physical violence, bullying on campus and bullying electronically loaded satisfactorily onto the ‘victimisation’ construct. Men were more likely to be victimised than women (p=0.002). Further, victimisation was positively associated with physical fights (path coefficient 0.37, 95% CI [0.16, 0.58], p<0.001). In turn, physical fights were positively associated with depressive symptoms (path coefficient 0.26, 95% CI [0.09, 0.42], p=0.003), and depressive symptoms were positively associated with suicidal ideation and attempts (path coefficient 0.95, 95% CI [0.59, 1.30], p<0.001). Victimisation had little direct effect on depressive symptoms (path coefficient 0.01, 95% CI [−0.19, 0.21], p=0.891) or suicidal ideation and attempts (path coefficient −0.06, 95% CI [−0.38, 0.27], p=0.739). Global fit measures were: chi-squared p-value 0.026, RMSEA=0.034, SRMR=0.179, CFI=0.960, and TLI=0.940.

**Table 3:** Path coefficients and covariance for the specified structural equation models, with and without ‘sexual abuse’ as an exogenous variable.

|  | Model with ‘sexual abuse’ |  | Model without ‘sexual abuse’ |  |
| --- | --- | --- | --- | --- |
|  | Parameter estimate<br>(95% CI) | p-value | Parameter estimate<br>(95% CI) | p-value |
| **Indicators of victimisation** |  |  |  |  |
| Physical violence during dating | 0.61 (0.29, 0.92) | <0.001 | 0.61 (0.29, 0.92) | <0.001 |
| Bullied on campus | 1.43 (0.44, 2.41) | 0.004 | 1.43 (0.44, 2.41) | 0.004 |
| Bullied electronically | 1.25 (0.50, 2.00) | 0.001 | 1.25 (0.50, 2.00) | 0.001 |
| **Regression for victimisation** |  |  |  |  |
| Age* | 0.01 (−0.09, 0.10) | 0.888 | 0.01 (−0.09, 0.10) | 0.888 |
| Gender** | −0.37 (−0.60, −0.13) | 0.002 | −0.37 (−0.60, −0.13) | 0.002 |
| **Regression for sexual abuse** |  |  |  |  |
| Age* | 0.04 (−0.25, 0.32) | 0.799 | — | — |
| Gender** | −0.09 (−0.60, 0.41) | 0.717 | — | — |
| **Regression for physical fight** |  |  |  |  |
| Victimisation | 0.37 (0.16, 0.58) | <0.001 | 0.37 (0.16, 0.57) | <0.001 |
| Sexual abuse | 0.08 (−0.20, 0.36) | 0.569 | — | — |
| Age* | 0.03 (−0.06, 0.13) | 0.505 | 0.04 (−0.06, 0.13) | 0.452 |
| Gender** | −0.11 (−0.36, 0.13) | 0.372 | −0.12 (−0.36, 0.12) | 0.337 |
| **Regression for depressive symptoms** |  |  |  |  |
| Victimisation | 0.01 (−0.19, 0.21) | 0.891 | 0.01 (−0.19, 0.21) | 0.927 |
| Sexual abuse | 0.10 (−0.18, 0.38) | 0.484 | — | — |
| Physical fight | 0.26 (0.09, 0.42) | 0.003 | 0.27 (0.10, 0.43) | 0.001 |
| Age* | 0.01 (−0.07, 0.10) | 0.759 | 0.02 (−0.06, 0.10) | 0.684 |
| Gender** | −0.12 (−0.35, 0.11) | 0.301 | −0.13 (−0.35, 0.09) | 0.259 |
| **Regression for suicidal ideation and attempts** |  |  |  |  |
| Victimisation | −0.06 (−0.38, 0.27) | 0.739 | −0.05 (−0.37, 0.26) | 0.740 |
| Sexual abuse | 0.17 (−0.24, 0.58) | 0.405 | — | — |
| Depressive symptoms | 0.95 (0.59, 1.30) | <0.001 | 0.96 (0.62, 1.30) | <0.001 |
| Age* | 0.06 (−0.14, 0.26) | 0.576 | 0.06 (−0.13, 0.26) | 0.523 |
| Gender** | 0.40 (−0.11, 0.90) | 0.122 | 0.38 (−0.11, 0.87) | 0.127 |

Path coefficients were standardised for the latent variable ‘victimisation’. \*For every one-year increase in age \*\* For women, as compared to men

When we simplified the model by removing the ‘sexual abuse’ variable, path coefficients were largely unchanged, but model fit improved. The simpler model had chi-squared p-value 0.036, RMSEA=0.034, SRMR=0.073, CFI=0.967, and TLI=0.956.

## Discussion

### Key findings

Our analysis of cross-sectional data for college students aged 18–22 years found satisfactory fit for our hypothesised structural equation model. Thus, the data support linkages of victimisation with adverse mental health outcomes. In particular, our model supports effects of victimisation on suicidal ideation and attempts, mediated by physical violence and depression. We also observed that men faced greater victimisation than women. In contrast to the effects of physical and bully-victimisation, we did not find strong effects of sexual abuse on any of the outcomes. We have considerable uncertainty about the ‘sexual abuse’ results, as the number of individuals reporting it was small.

### Victimisation and physical violence

We, like others, found that youth victimisation is positively associated with involvement in physical fights. A cross-sectional study among persons aged 15–19 in southern India found that bully-victimisation and violence experience were positively associated with risk factors for violence perpetration (Swain et al., 2014). Cross-sectional studies conducted in various cultures across the world corroborate these findings (Jeong et al., 2015; Cruz-Manrique et al., 2021; Ortiz-Marcos et al., 2022). Longitudinal evidence also shows that victimisation correlates with aggressive behaviour, such as psychological violence, cyberaggression and physical violence, at later time points in youth (Herrenkohl & Jung, 2016; Camacho et al., 2021; Costa et al., 2015). The psychological explanation for this linkage is “acting out”. Due to the inability to find mature and socially acceptable outlets, the individual externalises by engaging in violence, partly relieving the stress of victimisation (Meeus et al., 2016). Efforts to reduce violence among youth should therefore be twofold. First, youth environments, both online and offline, need to be made safer to prevent bullying and other forms of abuse. Second, youth need to be trained in coping strategies, so that they can deal effectively with stressors (Frank et al., 2014; Shelton et al., 2011).

### Victimisation and depression

Our model supports the hypothesis that recent victimisation predisposes to depression among adolescents and young adults. The true magnitude of the association could be larger than what we observed, because of selection bias. This is because we included only college attendees (not all enrolled students) as participants. Both victimisation and depression are known risk factors for school absenteeism (Brady et al., 2021; John et al., 2022). Thus, individuals with both victimisation and depression would have been underrepresented in our sample, diluting the observed association.

Longitudinal studies among youth show that the victimisation–depression relationship holds mostly in the short term, up to about two years (Fahy et al., 2016; Hemphill et al., 2015; Maurya et al., 2022; Sweeting et al., 2006). Though a study among Canadian adolescents di find lingering effects of victimisation on internalising symptoms, up to about eight years (Leadbeater et al., 2014), most other studies have found that these effects do not persist in the long run. However, within this short time frame, depression can have major consequences such as impairment of daily functioning and suicide attempts. Thus, the months following episodes of victimisation provide a window of opportunity, where the progression to depression can be stopped by appropriate cognitive and behavioural interventions.

Several cross-sectional studies have attempted to delineate pathways by which victimisation leads to depression. In addition to direct effects, researchers have found several mediators for this relationship: personal identity (Van Hoof et al., 2008), family self-concept (Cruz-Manrique et al., 2021), resilience (Bravo-Sanzana et al., 2023), and self-efficacy (Maurya et al., 2023). Though differently named, all these constructs are tied by a common thread: they represent psychological resources that an individual has at their disposal, to deal with stressful situations. In other words, victimisation increases an individual’s risk of depression not just directly, but also by exhausting their ability to endure stressors. Our finding that involvement in physical fights mediates the victimisation–depression relationship probably reflects this exhaustion of mental reserve.

### Physical violence and depression

Like our study, a preponderance of cross-sectional data support a positive association between violence perpetration and depression (Beckwith et al., 2022; Cohen et al., 2022; Tarriño-Concejero et al., 2023). However, the temporal order of this relationship is contested. For example, a seven-year prospective study among adolescents in Canada found support for the hypothesis that physical and relational aggression predict future depression. The reverse model did not fit the data as well (Blain-Arcaro & Vaillancourt, 2017). Similarly, peer aggression among Australian adolescents at age 14 was found to be positively associated with depression at age 17 (Moore et al., 2014). Considering these findings, we specified a path leading from ‘physical fight’ to ‘depressive symptoms’ in our model. The likely explanation is that the onset of depression follows an orderly pattern, where the individual first tries to cope with stressors by acting out, but once this fails, internalisation sets in (Blain-Arcaro & Vaillancourt, 2017). If we accept this interpretation, the period of physical aggression provides yet another window of opportunity for preventing depression.

Nevertheless, we recognise that the reverse is also plausible. A longitudinal study among junior high school students in China found that psychological distress (including depression) was positively associated with violent behaviour 10 months later, but the reverse was not true (Chen et al., 2023). In our analysis, reversing the order of the physical fight–depression relationship would result in a model that would be similar in terms of statistical fit, but very different in terms of implications. Such a model would imply that involvement in physical fights represents a consequence of depression, requiring specific assessment and management.

Finally, Jun, et al. (2015) offer a completely different explanation. Using data from a six-year prospective cohort study among adolescents in Chicago, USA, they argue that internalising and externalising symptoms correlate at a given point of time, but do not influence each other at subsequent time points (Jun et al., 2015). Such a model would suggest that some common and transient factor, such as an aggravating life event, is responsible for both aggression and depressive symptoms, and both resolve once this factor resolves.

### Victimisation and suicide-related behaviours

We noted a positive association between victimisation and suicidal ideation and attempts. This association should be interpreted subject to the same considerations of selection bias that we noted above for victimisation and depression. Also, previous work has shown that gender modifies the effect of victimisation on suicidal ideation. The effect of victimisation on suicidal ideation is stronger in girls than in boys (Nuñez-Fadda et al., 2022). Given the overrepresentation of women in our sample, our estimate of the association between victimisation and suicidal ideation might be upward-biased.

Still, our study replicates findings from previous cross-sectional studies across the globe. A pooled analysis of data from adolescents aged 12–17 years in 82 countries found that bullying was associated with over 25% increase in the prevalence odds of suicidal ideation (Biswas et al., 2020). Several cross-sectional studies have also attempted to understand mechanisms by which youth victimisation predisposes to suicidal behaviour (Cruz-Manrique et al., 2021; Hasan et al., 2021; Reed et al., 2015; Sutter et al., 2023). Consistent with our findings, a systematic review of studies among adolescents and young adults found that violent behaviour and depression mediate the link between victimisation and suicide. The evidence was less consistent regarding the role of substance use, self-compassion and psychological security as mediators(Khaki et al., 2022). Our analysis did not find a significant direct effect of victimisation on suicidal ideation and attempts, suggesting that these constructs, by themselves, are less important mediators than physical aggression and depression.

Interestingly, these findings are not uniformly reflected in longitudinal studies. While most cohort and case–control studies in the USA and Europe show positive associations of victimisation with suicide (Castellví et al., 2017; Miranda-Mendizabal et al., 2019), results from India are divergent. The UDAYA study, a large, prospective cohort study among persons aged 12–23 years in India, found that cyberbullying was positively associated with current suicidal ideation, but did not predict suicidal ideation three years later (Maurya et al., 2022). This shows differences in young persons’ coping abilities in India, probably due to the socio-cultural milieu, as family support for young people in India continues even into adulthood. Further prospective studies are required to accurately determine the long-term effect of youth victimisation on suicide in India, and identify the mediators in this causal path.

### Gender differences

One of our surprising findings was the higher reporting of dating violence by men. We could not find literature on dating violence amongst adolescents and young adults in this cultural context. In the United States, dating violence is experienced more commonly by the female gender (Cheung et al., 2023). We believe that there was some underreporting by women in our study; they might have been less comfortable than men in disclosing trauma inflicted by romantic partners. We also noted that men reported bullying more commonly than women. This applied for both traditional bullying and cyberbullying, and is in line with prior literature from India (Swain et al., 2014). Similarly, our finding of men being more involved in physical fights echoes prior literature (Swain et al., 2014). This shows that bullying and violence prevention programmes in the Indian scenario must be gender-sensitive, and give due attention to the needs of men.

Given our study size, we could not perform multi-group structural equation modelling for women versus men, or for women-only versus co-educational colleges. Rather, we calculated path coefficients adjusted for gender. Future research should also consider gender as a moderator of paths from victimisation to adverse mental health outcomes (Miranda-Mendizabal et al., 2019; Nuñez-Fadda et al., 2022; Sutter et al., 2023).

### Strengths

The strengths of our study were selection of participants using random sampling, use of a standardised tool, and adequate sample size. The anonymous, self-administered data collection meant that we could capture sensitive information, while minimising privacy concerns. Structural equation modelling allowed us to test a more complex hypothesis than would be possible with basic methods, such as regression for a single outcome variable. Therefore, we were able to discern linkages between multiple important constructs while adjusting for potential confounders. We specified our model based on current theory, thereby reducing the possibility of estimation errors due to model misspecification.

### Limitations

Yet, our work has its limitations, due to which results must be interpreted with caution. Firstly, this analysis was not specified while planning the study. Secondly, the data were collected in 2018, and prevalence of depression, suicide, etc. among college students could have changed since then. However, we consider it less likely that path coefficients would change over time. Thirdly, the design was cross-sectional. Hence, we could not comment on the temporal order in which events occurred. Also, recall bias could have occurred. For example, individuals with depression at the time of survey might have been more likely to recall victimisation experiences, because of their generally negative worldview. These problems occur with most cross-sectional studies. However, where evidence is limited and resource constraints do not permit longitudinal study designs, cross-sectional studies can play an important role in generating interesting hypotheses. Fourthly, our depression measure consisted of one question on sadness or hopelessness, and could have been inadequate to capture other aspects of depression, such as anhedonia and worthlessness. Fifthly, the recruitment of only college attendees could lead to selection bias.

### Implications

Though future longitudinal studies could provide additional insights, even at this stage, our study points to actionable areas for youth mental health in India. While life skills education can improve coping and can be readily incorporated into college curricula, (Tiwari et al., 2020) it is also important to tackle the root causes of violence, depression and suicide among youth. For this, stringent bullying prevention measures need to be implemented in colleges. Also, our study highlights the importance of cyberbullying in adverse mental health outcomes. Given the increasing numbers of Indian youth with smartphone and internet access, it is crucial to regulate social media and messaging platforms to minimise psychological ill-effects, while retaining the benefits of rapid communication. Early diagnosis and treatment of depression may help prevent progression to suicide. Accordingly, higher education authorities must ensure that mental health services provided by trained professionals are available and accessible to college students.

## Supporting information

Codebook and R script for analysis

TOP checklist for PCI Psychology

## Author contributions

Adhish Kumar Sethi: statistical analysis, writing first draft, editing draft.

PVM Lakshmi: conceptualisation, statistical analysis, reviewing draft.

Vikas Kumar Bhatia: conceptualisation, data collection, statistical analysis

Shubh Mohan Singh: conceptualisation, reviewing draft.

All authors read and approved the final version of the manuscript for submission.

## Acknowledgements

We thank the Department of Education (Chandigarh) for permission to conduct this study in colleges. We thank the principals of the colleges for their support, and the students for their participation in the study.

## Data availability statement

The data that support the findings of this study are publicly available (https://doi.org/10.1371/journal.pone.0340072.s005). The codebook and statistical analysis codes are provided as supplementary material with this article.

## Supplementary material

**Supplementary Figure 1:**
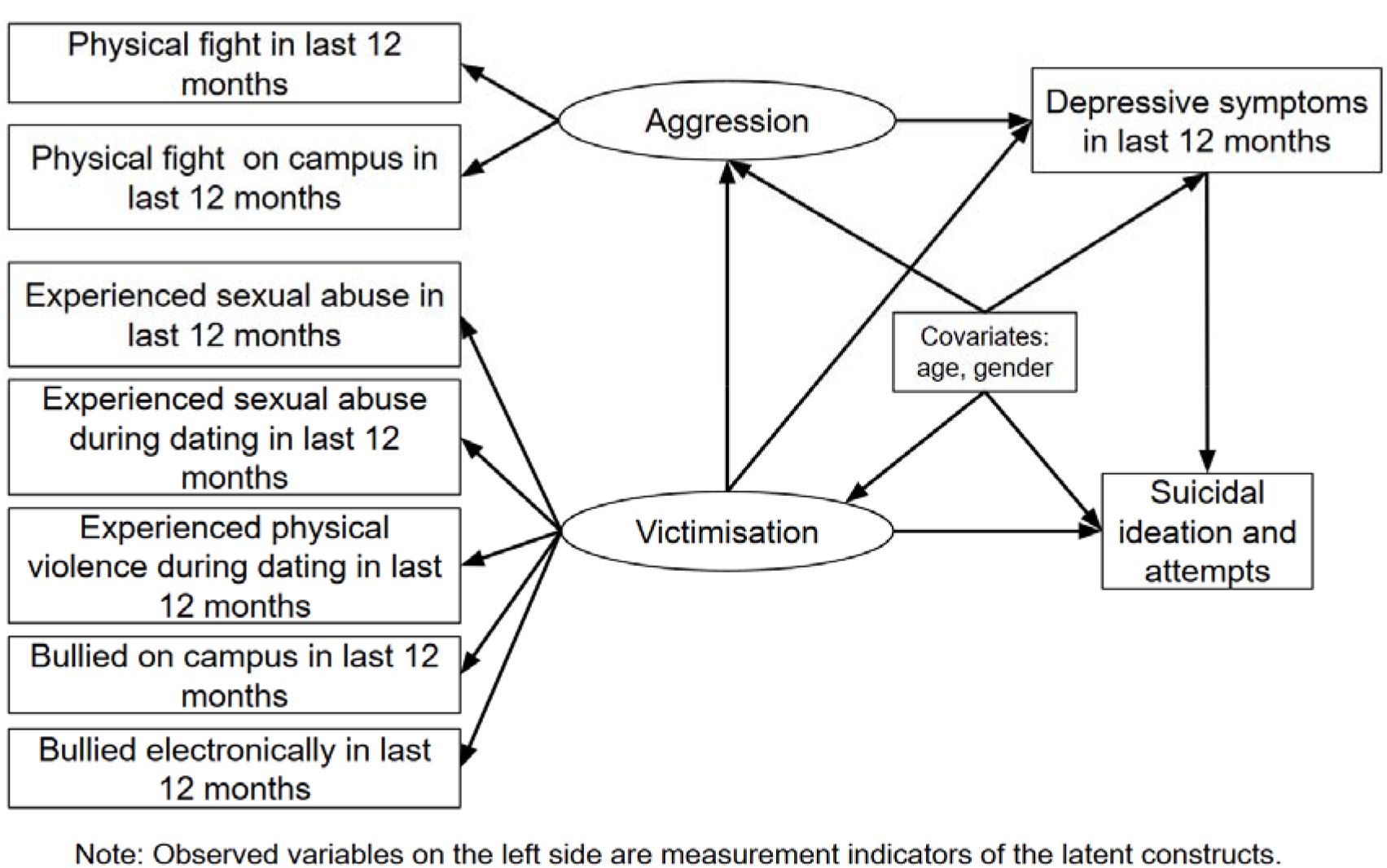
Hypothesised relationships among vicyimisation, depressive symptoms, suicide and physical violence among youth, based on previous literature.

**Supplementary Appendix 1:**
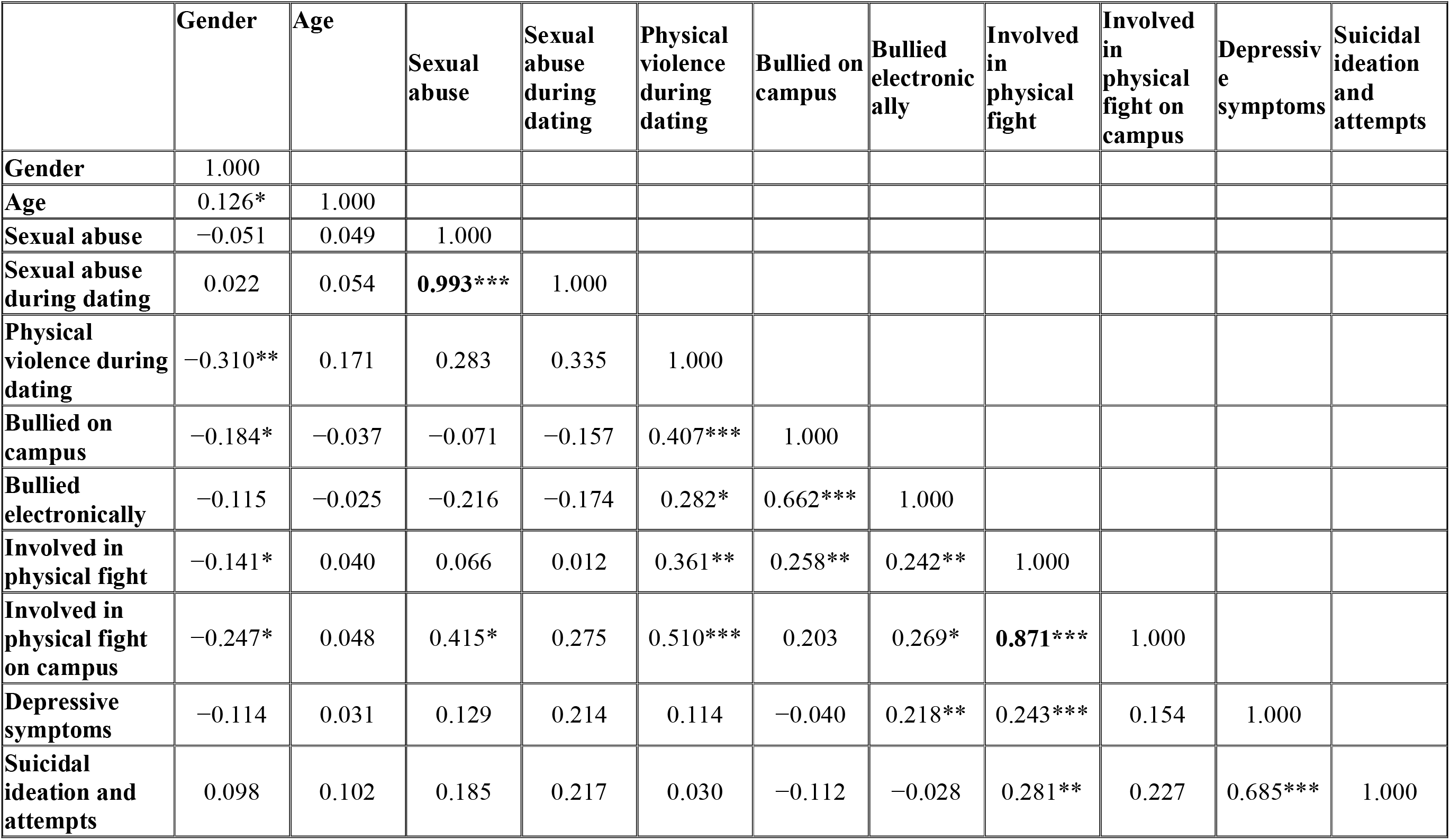
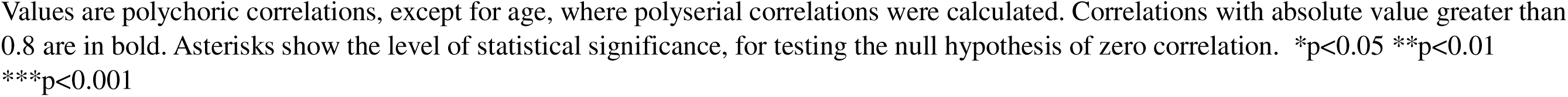
Correlation matrix of study variables.

|  | Gender | Age | Sexual abuse | Sexual abuse during dating | Physical violence during dating | Bullied on campus | Bullied electronically | Involved in physical fight | Involved in physical fight on campus | Depressive symptoms | Suicidal ideation and attempts |
| --- | --- | --- | --- | --- | --- | --- | --- | --- | --- | --- | --- |
| **Gender** | 1.000 |  |  |  |  |  |  |  |  |  |  |
| **Age** | 0.126* | 1.000 |  |  |  |  |  |  |  |  |  |
| **Sexual abuse** | -0.051 | 0.049 | 1.000 |  |  |  |  |  |  |  |  |
| **Sexual abuse during dating** | 0.022 | 0.054 | **0.993\*\*\*** | 1.000 |  |  |  |  |  |  |  |
| **Physical violence during dating** | -0.310** | 0.171 | 0.283 | 0.335 | 1.000 |  |  |  |  |  |  |
| **Bullied on campus** | -0.184* | -0.037 | -0.071 | -0.157 | 0.407*** | 1.000 |  |  |  |  |  |
| **Bullied electronically** | -0.115 | -0.025 | -0.216 | -0.174 | 0.282* | 0.662*** | 1.000 |  |  |  |  |
| **Involved in physical fight** | -0.141* | 0.040 | 0.066 | 0.012 | 0.361** | 0.258** | 0.242** | 1.000 |  |  |  |
| **Involved in physical fight on campus** | -0.247* | 0.048 | 0.415* | 0.275 | 0.510*** | 0.203 | 0.269* | **0.871\*\*\*** | 1.000 |  |  |
| **Depressive symptoms** | -0.114 | 0.031 | 0.129 | 0.214 | 0.114 | -0.040 | 0.218** | 0.243*** | 0.154 | 1.000 |  |
| **Suicidal ideation and attempts** | 0.098 | 0.102 | 0.185 | 0.217 | 0.030 | -0.112 | -0.028 | 0.281** | 0.227 | 0.685*** | 1.000 |

Values are polychoric correlations, except for age, where polyserial correlations were calculated. Correlations with absolute value greater than 0.8 are in bold. Asterisks show the level of statistical significance, for testing the null hypothesis of zero correlation. \* $p < 0.05$ \*\* $p < 0.01$ \*\*\* $p < 0.001$

