## Supplementary material for "Victimisation, depression and suicidal ideation: understanding interlinkages among youth in North India through structural equation modelling": Codebook and R script for analysis: Chandigarh youth risk behaviour codebook.docx

**Coding of study variables**

**Socio-demographics and self-reported weight and height**

| **Variable** | **Variable name** | **Variable type** | **Coding** |
| --- | --- | --- | --- |
| Serial number | sno | Quantitative, discrete | — |
| Age in completed years | age | Quantitative, discrete | — |
| Gender | gender | Categorical | 1 Male  2 Female  3 Transgender |
| Self-reported height in metres | ht | Quantitative, continuous | — |
| Self-reported weight in kg | wt | Quantitative, continuous | **—** |

**Safety**

| **Variable** | **Variable name** | **Variable type** | **Coding** |
| --- | --- | --- | --- |
| Seat belt use, while travelling in a car driven by someone else | seatbelt | Categorical, ordinal | 1 Never  2 Rarely  3 Sometimes  4 Most of the time  5 Always |
| Number of vehicle rides in the past 30 days, where driver was someone else under the influence of alcohol | ddriveother | Categorical, ordinal | 1 0 times  2 1 time  3 2 or 3 times  4 4 or 5 times  5 6 or more times |
| Number of times in past 30 days where person drove under the influence of alcohol | ddriveself | Categorical | 1 0 times  2 1 time  3 2 or 3 times  4 4 or 5 times  5 6 or more times  6 I did not drive a car or other vehicle during the past 30 days |
| Number of days in past 30 days where person texted/emailed/called during driving | mdrive | Categorical | 1 0 days  2 1 or 2 days  3 3 to 5 days  4 6 to 9 days  5 10 to 19 days  6 20 to 29 days  7 All 30 days  8 I did not drive a car or other vehicle during the past 30 days |
| Number of times person was involved in physical fight in past 12 months | fight | Categorical, ordinal | 1 0 times  2 1 time  3 2 or 3 times  4 4 or 5 times  5 6 or 7 times  6 8 or 9 times  7 10 or 11 times  8 12 or more times |
| Number of times person was involved in physical fight on campus in past 12 months | fightcampus | Categorical, ordinal | 1 0 times  2 1 time  3 2 or 3 times  4 4 or 5 times  5 6 or 7 times  6 8 or 9 times  7 10 or 11 times  8 12 or more times |
| Number of times person was physically hurt while dating in past 12 months | hurtdating | Categorical | 1 0 times  2 1 time  3 2 or 3 times  4 4 or 5 times  5 6 or more times  6 I did not date or go out with anyone during the past 12 months |
| Number of times person faced sexual abuse while dating in past 12 months | sexabusedating | Categorical | 1 0 times  2 1 time  3 2 or 3 times  4 4 or 5 times  5 6 or more times  6 I did not date anyone during the past 12 months |
| Number of times person faced sexual abuse in past 12 months | sexabuse | Categorical, ordinal | 1 0 times  2 1 time  3 2 or 3 times  4 4 or 5 times  5 6 or more times |
| Bullied on campus in past 12 months | cbullied | Categorical, binary | 0 No  1 Yes |
| Electronically bullied in past 12 months | ebullied | Categorical, binary | 0 No  1 Yes |
| Felt sad or hopeless for two weeks or more in past 12 months | sad | Categorical, binary | 0 No  1 Yes |
| Seriously considered suicide in past 12 months | csui | Categorical, binary | 0 No  1 Yes |
| Planned suicide in past 12 months | psui | Categorical | 0 No  1 Yes |
| Number of suicide attempts in past 12 months | asui | Categorical, ordinal | 1 0 times  2 1 time  3 2 or 3 times  4 4 or 5 times  5 6 or more times |
| Treatment by doctor or nurse after suicide attempt, in past 12 months | tsui | Categorical | 0 I did not attempt suicide during the past 12 months  1 Yes  2 No |

**Tobacco use**

| **Variable** | **Variable name** | **Variable type** | **Coding** |
| --- | --- | --- | --- |
| Ever smoked | smoke | Categorical, binary | 0 No  1 Yes |
| Age of initiation of smoking | agesmoke | Categorical | 1 10 years old or younger  2 10 to 13 years old  3 13 to 17 years old  4 17 years old or older  5 I have never tried cigarette smoking. |
| Number of days smoked in past 30 days | smokedays | Categorical, ordinal | 1 0 days  2 1 or 2 days  3 3 to 5 days  4 6 to 9 days  5 10 to 19 days  6 20 to 29 days  7 All 30 days |
| Number of cigarettes smoked each day, on days smoking was done in past 30 days | cigperday | Categorical | 1 Less than 1 cigarette per day  2 1 cigarette per day  3 2 to 5 cigarettes per day  4 6 to 10 cigarettes perday  5 11 to 20 cigarettes per day  6 More than 20 cigarettes per day  7 I did not smoke cigarettes during the past 30 days |
| Ever used an electronic vapour product (e-cigarettes, e-cigars, e-pipes, e-hookahs and hookah pens) | ends | Categorical, binary | 0 No  1 Yes |
| Ever used a smokeless tobacco product | smokeless | Categorical, binary | 0 No  1 Yes |
| Number of days used smokeless tobacco in past 30 days | smokelessdays | Categorical, ordinal | 1 0 days  2 1 or 2 days  3 3 to 5 days  4 6 to 9 days  5 10 to 19 days  6 20 to 29 days  7 All 30 days |
| Number of packets of smokeless tobacco used each day, in past 30 days | packperday | Categorical, ordinal | 1 Less than 1 packet.  2 1 packet per day.  3 2-5 packets per day.  4 5-10 packets per day.  5 More than 10 packets per day.  6 I did not use them |
| Tobacco quit attempts in past 12 months | tobquit | Categorical | 0 No  1 Yes  2 I did not use any tobacco products during the past 12months |

**Alcohol use**

| **Variable** | **Variable name** | **Variable type** | **Coding** |
| --- | --- | --- | --- |
| Ever used alcohol | alc | Categorical, binary | 0 No  1 Yes |
| Age of starting alcohol use | agealc | Categorical | 1 10 years old or younger  2 10 to 13 years old  3 13 to 17 years old  4 17 years old or older  5 I have never had a drink of alcohol other than a few sips |
| Number of days of alcohol use in past 30 days | alcdays | Categorical, ordinal | 1 0 days  2 1 or 2 days  3 3 to 5 days  4 6 to 9 days  5 10 to 19 days  6 20 to 29 days  7 All 30 days |
| Means of obtaining alcohol in past 30 days | alcmeans | Categorical | 1 Liquor store  2 Restaurant, bar, or club  3 Public event  4 Bought through someone else  5 Someone gave it  6 Store or family member  7 Others  8 I did not drink alcohol in last 30 days |
| Largest number of alcoholic drinks consumed in a row, in past 30 days | bingedrinks | Categorical | 1 1 or 2 drinks  2 3 drinks  3 4 drinks  4 5 drinks  5 6 or 7 drinks  6 8 or 9 drinks  7 10 or more drinks  8 I did not drink alcohol during the past 30 days |

**Substance use**

| **Variable** | **Variable name** | **Variable type** | **Coding** |
| --- | --- | --- | --- |
| Ever used cannabis | cann | Categorical, binary | 0 No  1 Yes |
| Age of starting cannabis use | agecann | Categorical | 1 8 years old or younger  2 9 or 10 years old  3 11 or 12 years old  4 13 or 14 years old  5 15 or 16 years old  6 17 years old or older  7 Never tried |
| Number of times used cannabis in life | cannlife | Categorical, ordinal | 1 0 times  2 1 or 2 times  3 3 to 9 times  4 10 to 19 times  5 20 to 39 times  6 40 or more times |
| Number of times used cannabis in past 30 days | cannmonth | Categorical, ordinal | 1 0 times  2 1 or 2 times  3 3 to 9 times  4 10 to 19 times  5 20 to 39 times  6 40 or more times |
| Ever used substance other than cannabis | other | Categorical, binary | 0 No  1 Yes |
| Number of times used substances other than cannabis in lifetime | otherlife | Categorical, ordinal | 1 0 times  2 1 or 2 times  3 3 to 9 times  4 10 to 19 times  5 20 to 39 times  6 40 or more times |
| Number of times used volatile solvents in lifetime | solvent | Categorical, ordinal | 1 0 times  2 1 or 2 times  3 3 to 9 times  4 10 to 19 times  5 20 to 39 times |
| Number of times used heroin during lifetime | heroin | Categorical, ordinal | 1 0 times  2 1 or 2 times  3 3 to 9 times  4 10 to 19 times  5 20 to 39 times  6 40 or more times |
| Number of times used MDMA/ecstasy in life | mdma | Categorical, ordinal | 1 0 times  2 1 or 2 times  3 3 to 9 times  4 10 to 19 times  5 20 to 39 times  6 40 or more times |
| Number of times in lifetime used prescription pain/cough medicine without doctor’s advice | med | Categorical, ordinal | 1 0 times  2 1 or 2 times  3 3 to 9 times  4 10 to 19 times  5 20 to 39 times  6 40 or more times |
| Number of times used steroids without doctor’s advice | steroid | Categorical, ordinal | 1 0 times  2 1 or 2 times  3 3 to 9 times  4 10 to 19 times  5 20 to 39 times  6 40 or more times |
| Number of times injected drugs in lifetime | inj | Categorical, ordinal | 1 0 times  2 1 time  3 2 or more times |
| Offered, sold, or given illegal drug on or around college property in past 12 months | illegal | Categorical, binary | 0 No  1 Yes |

**Diet and nutrition**

| **Variable** | **Variable name** | **Variable type** | **Coding** |
| --- | --- | --- | --- |
| Perceived weight | percwt | Categorical, ordinal | 1 Very underweight  2 Slightly underweight  3 About the right weight  4 Slightly overweight  5 Very overweight |
| Efforts to change weight | effortwt | Categorical | 1 Lose weight  2 Gain weight  3 Stay the same weight  4 I am not trying anything |
| Number of times drank fruit juice in past seven days | juice | Categorical | 1 1 to 3 times during the past 7 days  2 4 to 6 times during the past 7 days  3 1 time per day  4 2 times per day  5 3 times per day  6 4 or more times per day  7 I did not drink 100% fruit juice during the past 7 days |
| Number of times fruit was consumed in past seven days | fruit | Categorical | 1 1 to 3 times during the past 7 days  2 4 to 6 times during the past 7 days  3 1 time per day  4 2 times per day  5 3 times per day  6 4 or more times per day  7 I did not eat fruit during the past 7 days |
| Number of times potato was consumed in past seven days | pot | Categorical, ordinal | 1 1 to 3 times during the past 7 days  2 4 to 6 times during the past 7 days  3 1 time per day  4 2 times per day  5 3 times per day  6 4 or more times per day  7 I did not eat potatoes during the past 7 days |
| Number of times vegetables were consumed in past seven days | veg | Categorical, ordinal | 1 1 to 3 times during the past 7 days  2 4 to 6 times during the past 7 days  3 1 time per day  4 2 times per day  5 3 times per day  6 4 or more times per day  7 I did not eat other vegetables during the past 7 days |
| Number of times carbonated drinks were consumed in past seven days | fizzy | Categorical, ordinal | 1 1 to 3 times during the past 7 days  2 4 to 6 times during the past 7 days  3 1 time per day  4 2 times per day  5 3 times per day  6 4 or more times per day  7 I did not drink soda or pop during the past 7 days |
| Number of glasses of milk consumed in past seven days | milk | Categorical | 1 1 to 3 glasses during the past 7 days  2 4 to 6 glasses during the past 7 days  3 1 glass per day  4 2 glasses per day  5 3 glasses per day  6 4 or more glasses per day  7 I did not drink milk during the past 7 days |
| Number of days breakfast was consumed in past seven days | break | Quantitative, discrete | — |

**Physical activity and sleep**

| **Variable** | **Variable name** | **Variable type** | **Coding** |
| --- | --- | --- | --- |
| Number of days with at least 60 minutes of physical activity in past seven days | pa | Quantitative, discrete | — |
| Number of days in past seven days on which muscle strengthening exercise was done | muscle | Quantitative, discrete | — |
| Number of hours spent watching TV on average school day | tv | Categorical | 1 Less than 1 hour per day  2 1 hour per day  3 2 hours per day  4 3 hours per day  5 4 hours per day  6 5 or more hours per day  7 I do not watch TV on an average school day |
| Hours spent using computer for non-school work on average school day | comp | Categorical | 1 Less than 1 hour per day  2 1 hour per day  3 2 hours per day  4 3 hours per day  5 4 hours per day  6 5 or more hours per day  7 I do not use a computer other than for school assignment. |
| Number of sports teams on which played in past 12 months | team | Categorical, ordinal | 1 0 teams  2 1 team  3 2 teams  4 3 or more teams |
| Duration of sleep on an average school night | sleep | Categorical, ordinal | 1 4 or less hours  2 5 hours  3 6 hours  4 7 hours  5 8 hours  6 9 hours  7 10 or more hours |
| Grades in school over past 12 months | grade | Categorical | 1 Mostly A's  2 Mostly B's  3 Mostly C's  4 Mostly D's  5 Mostly F's  6 None of these grades  7 Not sure |
| Cognitive difficulty | cogdiff | Categorical, binary | 0 No  1 Yes |

**Sexual Behaviour**

| **Variable** | **Variable name** | **Variable type** | **Coding** |
| --- | --- | --- | --- |
| Ever had sexual intercourse | sex | Categorical, binary | 0 No  1 Yes |
| Age at first sexual intercourse | agesex | Categorical | 1 11 years old or younger  2 12 years old  3 13 years old  4 14 years old  5 15 years old  6 16 years old  7 17 years old or older  8 Never had sexual intercourse |
| Number of sexual partners in lifetime | partner | Categorical | 1 1 person  2 2 people  3 3 people  4 4 people  5 5 people  6 6 or more people  7 I have never had sexual intercourse |
| Number of sexual partners in past three months | partner3m | Categorical | 1 I have had sexual intercourse, but not during the past 3 months  2 1 person  3 2 people  4 3 people  5 4 people  6 5 or more people  7 I have never had sexual intercourse |
| Alcohol/drug use before last sex | subsex | Categorical | 0 No  1 Yes  2 Never had sex |
| Condom use at last sexual intercourse | condom | Categorical | 0 No  1 Yes  2 Never had sex |
| Contraceptive used at last sex | contra | Categorical | 1 No method was used to prevent pregnancy  2 Birth control pills  3 Condoms  4 Withdrawal or some other method  5 Not sure  6 I have never had sexual intercourse |
| Gender of sexual contacts during lifetime | gendercont | Categorical | 1 Females  2 Males  3 Females and males  4 I have never had sexual contact |
| Sexual orientation | orient | Categorical | 1 Heterosexual (straight)  2 Gay or lesbian  3 Bisexual  4 Not sure |

**Recoded/transformed variables**

| **Variable** | **Variable name** | **Variable type** | **Coding** |
| --- | --- | --- | --- |
| BMI in kg/m^2^ | bmi | Quantitative, continuous | — |
