## Supplementary material for "Victimisation, depression and suicidal ideation: understanding interlinkages among youth in North India through structural equation modelling": Codebook and R script for analysis: Mental health SEM script.R

```
# SEM script

# Load the package
library(lavaan)

# First import data as a data frame called "d".

# Recoding variables

# Making an ordinal variable for suicide:
d$sui <- ifelse(d$csui==0, 1, 
		ifelse(d$psui==0, 2,
		ifelse(d$asui==1, 3, 
		ifelse(d$tsui==2, 4, 5))))

# Recoding other ordinal variables as binary (1=no, 2=yes)
d$cbullied2<-d$cbullied+1 # Faced bullying in the campus 
d$ebullied2<-d$ebullied+1 # Faced cyber bullying 
d$sad2<-d$sad+1 # Felt sad or hopeless
d$hurtdating2 <- (d$hurtdating>1 & d$hurtdating<6)+1
d$sexabuse2 <- ifelse(d$sexabuse==1, 1, 2)
d$sexabusedating2 <- (d$sexabusedating>1 & d$sexabusedating<6)+1
d$fight2 <- (d$fight>1)+1
d$fightcampus2 <- ifelse(d$fightcampus==1, 1, 2)

# Summarising variables
# Gender
table(d$gender, useNA='ifany')
round(100*prop.table(table(d$gender, useNA='ifany')), 1)
# Age
summary(d$age[d$gender==1]); summary(d$age[d$gender==2]); summary(d$age)
# Sexual abuse: binary
addmargins(table(d$sexabuse2, d$gender, useNA='ifany'))
round(100*prop.table(addmargins(table(d$sexabuse2, d$gender, useNA='ifany'),
		margin=2), margin=2), 1)
# Sexual abuse: disaggregated by frequency
addmargins(table(d$sexabuse, d$gender, useNA='ifany'))
# Sexual abuse during dating: binary
addmargins(table(d$sexabusedating2, d$gender, useNA='ifany'))
round(100*prop.table(addmargins(table(d$sexabusedating2, d$gender, useNA='ifany'),
		margin=2), margin=2), 1)
# Sexual abuse during dating: disaggregated by frequency
addmargins(table(d$sexabusedating, d$gender, useNA='ifany'))
# Physical violence during dating: binary
addmargins(table(d$hurtdating2, d$gender, useNA='ifany'))
round(100*prop.table(addmargins(table(d$hurtdating2, d$gender, useNA='ifany'),
		margin=2), margin=2), 1)
# Physical violence during dating: disaggregated by frequency
addmargins(table(d$hurtdating, d$gender, useNA='ifany'))
# Bullied on campus in last 12 months
addmargins(table(d$cbullied2, d$gender, useNA='ifany'))
round(100*prop.table(addmargins(table(d$cbullied2, d$gender, useNA='ifany'),
		margin=2), margin=2), 1)
# Bullied electronically in last 12 months
addmargins(table(d$ebullied2, d$gender, useNA='ifany'))
round(100*prop.table(addmargins(table(d$ebullied2, d$gender, useNA='ifany'),
		margin=2), margin=2), 1)
# Fight in last 12 months: binary
addmargins(table(d$fight2, d$gender, useNA='ifany'))
round(100*prop.table(addmargins(table(d$fight2, d$gender, useNA='ifany'),
		margin=2), margin=2), 1)
# Fight in last 12 months: disaggregated by frequency
addmargins(table(d$fight, d$gender, useNA='ifany'))
# Fight on campus in last 12 months: binary
addmargins(table(d$fightcampus2, d$gender, useNA='ifany'))
round(100*prop.table(addmargins(table(d$fightcampus2, d$gender, useNA='ifany'),
		margin=2), margin=2), 1)
# Fight on campus in last 12 months: disaggregated by frequency
addmargins(table(d$fightcampus, d$gender, useNA='ifany'))
# Depressive symptoms last 12 months
addmargins(table(d$sad2, d$gender, useNA='ifany'))
round(100*prop.table(addmargins(table(d$sad2, d$gender, useNA='ifany'),
		margin=2), margin=2), 1)
# Suicide
addmargins(table(d$sui, d$gender, useNA='ifany'))
round(100*prop.table(addmargins(table(d$sui, d$gender, useNA='ifany'),
		margin=2), margin=2), 1)

# Polychoric correlations
cormatrix <- lavCor(d[,c("gender","age","sexabuse2","sexabusedating2", 
		"hurtdating2", "cbullied2", "ebullied2", "fight2", "fightcampus2", 
		"sad2", "sui")], 
	missing='pairwise', 
	ordered=c("gender","sexabuse2","sexabusedating2", "hurtdating2", "cbullied2",
		"ebullied2", "fight2", "fightcampus2", "sad2", "sui"),
	output="lavaan", estimator="two.step", se="standard")
summary(cormatrix)

# Factor analysis for the victimisation construct
# EFA

# One-factor model
efa1 <- efa(data=d[,c("hurtdating2","sexabuse2","sexabusedating2","cbullied","ebullied")],
	nfactors = 1, rotation = "geomin", missing="listwise",
	ordered=c("hurtdating2","sexabuse2","sexabusedating2","cbullied","ebullied"))
summary(efa1) # Heywood case! sexabuse2 has negative unique variance
fitMeasures(efa1, fit.measures=c("pvalue","rmsea","srmr","cfi","tli"))

# Two-factor model
efa2 <- efa(data=d[,c("hurtdating2","sexabuse2","sexabusedating2","cbullied","ebullied")],
	nfactors = 2, rotation = "geomin", missing="listwise",
	ordered=c("hurtdating2","sexabuse2","sexabusedating2","cbullied","ebullied"))
summary(efa2) # Heywood case! sexabusedating2 has negative unique variance
fitMeasures(efa2, fit.measures=c("pvalue","rmsea","srmr","cfi","tli"))

# Removing sexual abuse during dating
efa3 <- efa(data=d[,c("hurtdating2","sexabuse2","cbullied","ebullied")],
	nfactors = 1, rotation = "geomin", missing="listwise",
	ordered=c("hurtdating2","sexabuse2","cbullied","ebullied"))
summary(efa3, cutoff=0.001, pvalue=TRUE)
fitMeasures(efa3, fit.measures=c("pvalue","rmsea","srmr","cfi","tli"))
# So it's better to remove sexual abuse during dating
# Also, sexual abuse has a small, negative and uncertain factor loading.

# Removing sexual abuse entirely
efa4 <- efa(data=d[,c("hurtdating2","cbullied","ebullied")],
	nfactors = 1, rotation = "geomin", missing="listwise",
	ordered=c("hurtdating2","cbullied","ebullied"))
summary(efa4)
fitMeasures(efa4, fit.measures=c("pvalue","rmsea","srmr","cfi","tli"))


# CFA
cfa1 <-cfa(model='victim=~hurtdating2 + sexabuse2 + cbullied + ebullied',
	data=d, 
	ordered=c("hurtdating2","sexabuse2","cbullied","ebullied"))
summary(cfa1, standardized=TRUE)
fitMeasures(cfa1, fit.measures=c("pvalue","rmsea","srmr","cfi","tli"))

cfa2 <-cfa(model='victim=~hurtdating2 + cbullied + ebullied',
	data=d, 
	ordered=c("hurtdating2","cbullied","ebullied"))
summary(cfa2, standardized=TRUE)
fitMeasures(cfa2, fit.measures=c("pvalue","rmsea","srmr","cfi","tli"))

# Try with 'physical fight' as a victimisation indicator
cfa3 <-cfa(model='victim=~hurtdating2 + fight2 + cbullied + ebullied',
	data=d, 
	ordered=c("hurtdating2","fight2","cbullied","ebullied"))
summary(cfa3, standardized=TRUE)
fitMeasures(cfa3, fit.measures=c("pvalue","rmsea","srmr","cfi","tli"))
# Fit worsens. Also, literature supports physical fight as consequence rather
# than indicator.

# Multi-group CFA for CFA model 2; for measurement invariance
# Configural invariance
cfa2_config <- cfa(model='victim=~hurtdating2 + cbullied + ebullied', 
	d, ordered=c("hurtdating2","cbullied","ebullied"),
	group = "gender") # Heywood case!
# Weak (metric) invariance
cfa2_weak <- cfa(model='victim=~hurtdating2 + cbullied + ebullied', 
	d, ordered=c("hurtdating2","cbullied","ebullied"),
	group = "gender", group.equal = c("loadings"))
# Strong (scalar) invariance
cfa2_strong <- cfa(model='victim=~hurtdating2 + cbullied + ebullied', 
	d, ordered=c("hurtdating2","cbullied","ebullied"),
	group = "gender", group.equal = c("intercepts", "loadings"))
# Comparing CFA models
lavTestLRT(cfa2_config, cfa2_weak, cfa2_strong)

# Main SEM analysis
# Specifying a model
model1<-'
	! Latent variables
	victim=~hurtdating1 + sexabuse + sexabusedating1 + cbullied + ebullied
	aggr =~ fight + fightcampus
	
	! Regressions
	sad ~ age + gender + victim + aggr
	victim ~ age + gender
	aggr ~ victim + age + gender
	sui ~ sad + victim + age + gender
	'

# Fitting
sem1<-sem(model1, ordered=c('hurtdating1','sexabuse','sexabusedating1',
	'cbullied','ebullied','sad','sui','fight','fightcampus'),d,
	estimator="WLSMV", missing="pairwise", parameterization="theta")
# Convergence issues!
# Assessment 
summary(sem1)
parTable(sem1)[,c("lhs","op","rhs","free","user","est")]
fitMeasures(sem1, fit.measures=c("pvalue","rmsea","srmr","cfi","tli"))

# Simplifying, keeping binary variables
model2<-'
	! Latent variables
	victim=~hurtdating2 + sexabuse2 + sexabusedating2 + cbullied + ebullied
	aggr =~ fight2 + fightcampus2
	
	! Regressions
	sad ~ age + gender + victim + aggr
	victim ~ age + gender
	aggr ~ victim + age + gender
	sui ~ sad + victim + age + gender
	'
sem2<-sem(model2, ordered=c('hurtdating2','sexabuse2', 'sexabusedating2',
	'cbullied','ebullied','sad','sui','fight2','fightcampus2'),d,
	estimator="WLSMV", missing="pairwise", parameterization="theta")
# Assessment 
summary(sem2)
parTable(sem2)[,c("lhs","op","rhs","free","user","est")]
fitMeasures(sem2, fit.measures=c("pvalue","rmsea","srmr","cfi","tli"))
# Though this model converged, the estimates are weird and the fit is not great.

# Figuring out problems
table(d$fight2, d$fightcampus2) # Too few individuals with fight on campus
# Also recall the large correlation between fight2 and fightcampus2
table(d$sexabuse2, d$sexabusedating2) # Too few with sexual abuse
# Even among them, most had it during dating.


# Simplifying the model: removing 'sexabusedating2'
# Also removing the latent 'aggression' and just keeping 'fight2'
model3 <- '	
	! Latent variables
	victim=~hurtdating2 + sexabuse2 + cbullied + ebullied
	
	! Regressions
	sad ~ age + gender + victim + fight2
	victim ~ age + gender
	fight2 ~ victim + age + gender
	sui ~ sad + victim + age + gender
	'

# Fitting
sem3<-sem(model3, ordered=c('hurtdating2','sexabuse2',
	'cbullied','ebullied','sad','sui','fight2'),d,
	estimator="WLSMV", missing="pairwise", parameterization="theta")

# Assessment 
summary(sem3, standardized=T)
parTable(sem3)[,c("lhs","op","rhs","free","user","est")]
fitMeasures(sem3, fit.measures=c("pvalue","rmsea","srmr","cfi","tli"))
# Not bad. SRMR is quite high, though.

# Alternative: adding sexual abuse as another variable, 
# outside of victimisation, and uncorrelated with it.
model4 <- '	
	! Latent variables
	victim=~hurtdating2 + cbullied + ebullied
	
	! Regressions
	sad ~ gender + victim + sexabuse2 + age + fight2
	victim ~ gender + age
	sexabuse2 ~ gender + age
	fight2 ~ victim + sexabuse2 + gender + age
	sui ~ sad + victim + sexabuse2 + gender + age

	! Correlation
	victim~~0*sexabuse2
	'

# Fitting
sem4<-sem(model4, ordered=c('hurtdating2','sexabuse2',
	'cbullied','ebullied','sad','sui','fight2'),d,
	estimator="WLSMV", missing="pairwise", parameterization="theta")

# Assessment 
summary(sem4, standardized=TRUE)
standardizedSolution(sem4, type='std.lv')
parTable(sem4)[,c("lhs","op","rhs","free","user","est")]
fitMeasures(sem4, fit.measures=c("pvalue","rmsea","srmr","cfi","tli"))
# OK, but SRMR is still quite high and p-value quite low.

# Further simplifying, by removing sexual abuse entirely.
model5 <- '	
	! Latent variables
	victim=~hurtdating2 + cbullied + ebullied

	! Regressions
	sad ~ gender + victim + age + fight2
	victim ~ gender + age
	fight2 ~ victim + gender + age
	sui ~ sad + victim + gender + age
	'

# Fitting
sem5<-sem(model5, ordered=c('hurtdating2',
	'cbullied','ebullied','sad','sui','fight2'),d,
	estimator="WLSMV", missing="pairwise", parameterization="theta")

# Assessment 
summary(sem5, standardized=TRUE)
standardizedSolution(sem5, type='std.lv')
parTable(sem5)[,c("lhs","op","rhs","free","user","est")]
fitMeasures(sem5, fit.measures=c("pvalue","rmsea","srmr","cfi","tli"))
# Fit seems to have improved now. Retain this model.
lavResiduals(sem5)
# Important correlation residuals: fight2 and hurtdating2; sad and cbullied; 
# sad and ebullied; ebullied and hurtdating2
# Important residual slopes: hurtdating2 and gender; hurtdating2 and age

# Finding number who had missing data for ALL endogenous variables in models 4 and 5.
# Model 4
sum(is.na(d$sexabuse2) & is.na(d$hurtdating2) & is.na(d$cbullied2) 
	& is.na(d$ebullied2) & is.na(d$fight2) & is.na(d$sad2) & is.na(d$sui))
# Zero, so it must be zero for model 5 too.

# Trying to account for the residual covariances and slopes in model 5.
# Adding just a residual covariance between 'ebullied' and 'hurtdating2'.
model6 <- '	
	! Latent variables
	victim=~hurtdating2 + cbullied + ebullied

	! Regressions
	sad ~ gender + victim + age + fight2
	victim ~ gender + age
	fight2 ~ victim + gender + age
	sui ~ sad + victim + gender + age
	
	! Residual covariances
	ebullied ~~ hurtdating2
	'

# Fitting
sem6<-sem(model6, ordered=c('hurtdating2',
	'cbullied','ebullied','sad','sui','fight2'),d,
	estimator="WLSMV", missing="pairwise", parameterization="theta")
# Assessment
summary(sem6, standardized=TRUE)
standardizedSolution(sem6, type='std.lv')
parTable(sem6)[,c("lhs","op","rhs","free","user","est")]
fitMeasures(sem6, fit.measures=c("pvalue","rmsea","srmr","cfi","tli"))
lavResiduals(sem6)
# Though the addition improves fit, with minimal influence on other estimates, 
# it reduces parsimony, and is difficult to justify theoretically.
# Something similar happens on adding other covariances (e.g., sad ~~ cbullied)
# and slopes (e.g., hurtdating2 ~ gender).

# Considering victimisation as FORMED (rather than INDICATED) by the items.
model7 <- '	
	! Latent variables
	victim <~ 1*cbullied + ebullied + hurtdating2

	! Regressions
	sad ~ gender + victim + age + fight2
	cbullied ~ age + gender
	ebullied ~ age + gender
	hurtdating2 ~ age + gender
	fight2 ~ victim + gender + age
	sui ~ sad + victim + gender + age
	
	! Variances and covariances
	victim ~~ 1*victim
	hurtdating2 ~~ cbullied
	cbullied ~~ ebullied
	ebullied~~hurtdating'

# Fitting
sem7<-sem(model7, ordered=c('sad','sui','fight2','cbullied',
		'ebullied', 'hurtdating2'),d, 
	parameterization="theta", estimator="WLSMV")
# Assessment
summary(sem7, standardized=TRUE)
standardizedSolution(sem7, type='std.lv')
parTable(sem7)[,c("lhs","op","rhs","free","user","est")]
fitMeasures(sem7, fit.measures=c("pvalue","rmsea","srmr","cfi","tli"))
lavResiduals(sem7)
# Failure to fit model.

## Multi-group SEM: by gender
# Since we have
model4_multi <- '	
	! Latent variables
	victim=~hurtdating2 + cbullied + ebullied

	! Regressions
	sad ~ victim + age + fight2
	victim ~ age
	fight2 ~ victim + age
	sui ~ sad + victim + age
	'

# Fitting
# All parameters free
sem4a<-sem(model4_multi, ordered=c('hurtdating2',
	'cbullied','ebullied','sad','sui','fight2'),d,group="gender", 
	estimator="WLSMV", missing="pairwise", parameterization="theta")
# Equal factor loadings across groups
sem4b<-sem(model4_multi, ordered=c('hurtdating2',
	'cbullied','ebullied','sad','sui','fight2'),d,group="gender",
	group.equal="loadings", estimator="WLSMV", 
	missing="pairwise", parameterization="theta")
# Equal factor loadings and regressions
sem4c<-sem(model4_multi, ordered=c('hurtdating2',
	'cbullied','ebullied','sad','sui','fight2'),d,group="gender",
	group.equal=c("loadings","regressions"), estimator="WLSMV", 
	missing="pairwise", parameterization="theta")
# Comparing models
lavTestLRT(sem4a, sem4b, sem4c)
# The multi-group analysis has one major problem:
# The extremely high standard error for the 'electronic bullying' loading
# among women in Model 4a. This is likely due to small group sizes.
```
