## Supplementary material for "Victimisation, depression and suicidal ideation: understanding interlinkages among youth in North India through structural equation modelling": TOP checklist for PCI Psychology

This checklist must be completed and included as an appendix at the end of your preprint prior to submission to PCI Psychology. Manuscripts without this document included as an appendix will be returned to authors without review.

The policy of PCI Psychology is to recommend papers **only if the data, methods used in the analysis, and any digital materials used to conduct the research are clearly and precisely documented and are maximally available** to any researcher for purposes of reproducing the results or replicating the procedure. PCI Psych follows the principle of “as open as possible, as closed as necessary.” See the [PCI Psychology TOP Guidelines](https://psych.peercommunityin.org/help/help_generic#h_89713720928531739315150765) and the [Guide for Authors](https://psych.peercommunityin.org/help/guide_for_authors) for more details on policies and expectations.

**First author name (last/family, first/given):** Sethi, Adhish Kumar

**Preprint DOI or URL:** <https://doi.org/10.64898/2026.09.14.26362969>

**Section 1: Data**

**Does your manuscript contain reports of any data?**

☒ Yes (continue with next question)

☐ No (skip to Section 2):

**Are appropriately anonymised raw data available within a trusted digital repository?**

☒ Yes, available at this link: <https://doi.org/10.1371/journal.pone.0340072.s005>

☐ No, justification: Click or tap here to enter text.

**Are third-party data cited in the manuscript, with a DOI? (e.g., for preexisting data, data deposited in a repository; see** [**Data citation – A guide to best practice**](https://data.europa.eu/doi/10.2830/59387)**)**

☐ Yes, the DOI is as follows: Click or tap here to enter text.

☒ No, justification: No third-party data or pre-existing data were used. All raw data are available at the above link.

**Is there a data dictionary and/or readme file included with the data to make it interpretable?**

☒ Yes, available at this link: <https://doi.org/10.64898/2026.09.14.26362969>

☐ No, justification: Click or tap here to enter text.

**Do you indicate in the manuscript how the sample size was determined?**

☒ Yes.

☐ No, justification: Click or tap here to enter text.

**Do you report all data exclusions (e.g., outliers, careless responders)?**

☒ Yes.

☐ No, justification: Click or tap here to enter text.

**Do you report all inclusion/exclusion criteria and when they were established?**

☒ Yes.

☐ No, justification: Click or tap here to enter text.

**Are all measures, questions, and/or conditions used in the study described in the manuscript or available in the supplemental material?**

☒ Yes.

☐ No, justification: Click or tap here to enter text.

**Section 2: Analysis Scripts/Code/Codebooks**

**Does your manuscript contain any analysis of quantitative or qualitative data?**

☒ Yes (continue with next question)

☐ No (skip to Section 3):

**Are third-party analysis scripts/code (e.g., R, Stata), codebooks, or other relevant documentation available within a trusted digital repository?**

☒ Yes, available at this link: <https://doi.org/10.64898/2026.09.14.26362969>

☐ No, justification: Click or tap here to enter text.

**Are the analysis scripts/code (e.g., R, Stata), codebooks, or other relevant documentation cited in the manuscript, with a DOI?**

☒ Yes, the DOI is as follows: <https://doi.org/10.64898/2026.09.14.26362969>

☐ No, justification: Click or tap here to enter text.

**Section 3: Study Materials**

**Does your manuscript contain any research materials (e.g., stimuli, programming code, questionnaires, interview protocols)?**

☒ Yes (continue with next question)

☐ No (skip to Section 4):

**Are all study materials and descriptions of study procedures available within a trusted digital repository?**

☒ Yes, available at this link: <https://doi.org/10.1371/journal.pone.0340072.s001>

☐ No, justification: Click or tap here to enter text.

**Are all third-party study materials, descriptions of study procedures, or other relevant documents cited in the manuscript, with a DOI?**

☐ Yes, the DOI is as follows: Click or tap here to enter text.

☒ No, justification: We did not use third-party study material. All the material that we used is provided as supplementary material.

**Section 4: Preregistration**

**Were any aspects of your manuscripts preregistered?**

☐ Yes (continue with next question)

☒ No (do not complete the rest of the form):

**Does the manuscript contain an accessible link to the preregistration?**

☐ Yes, available at this link: Click or tap here to enter text.

☐ No, justification: Click or tap here to enter text.

**Do you clearly indicate in the manuscript which parts were preregistered and which parts were not?**

☐ Yes.

☐ No, justification: Click or tap here to enter text.

**Are all preregistered analyses reported in the text or linked in the supplemental material?**

☐ Yes.

☐ No, justification: Click or tap here to enter text.

**Are all deviations from the preregistration plan clearly disclosed in the manuscript (either in text or in a table)?**

☐ Yes.

☐ No, justification: Click or tap here to enter text.
